# Pathways and barriers to becoming physician-scientists for first-generation individuals

**DOI:** 10.1101/2024.03.17.24304448

**Authors:** Briana Christophers, Briana Macedo, Jessica Weng, Michael C. Granovetter, Rachit Kumar, Chynna Smith, Olaf S. Andersen, Catharine Boothroyd

**Affiliations:** Weill Cornell/Rockefeller/Sloan Kettering Tri-Institutional MD-PhD Program, New York, NY; Medical Scientist Training Program of the Perelman School of Medicine at the University of Pennsylvania, Philadelphia, PA; Mayo Clinic Medical Scientist Training Program, Rochester, MN 55905, USA; University of Pittsburgh-Carnegie Mellon University Medical Scientist Training Program, Pittsburgh, PA; Albert Einstein College of Medicine Medical Scientist Training Program, Bronx, NY

## Abstract

**Introduction:** Physician-scientists are uniquely positioned to contribute translational research that will impact patient care and our understanding of disease. Having a diverse cadre of physician-scientists is critical to ensuring that the biomedical research enterprise explores the breadth of problems affecting the nation’s health. The National Institutes of Health has identified diversity, including educational background, to be important for the biomedical workforce. In 2020, less than ten percent of MD-PhD program matriculants were the first in their families to pursue higher education (first-generation) despite the majority of the US population having less than a Bachelor’s degree. Little is known about the specific challenges that first-generation students face, which makes it challenging to address this gap in matriculation.

**Methods:** This qualitative study used a phenomenological approach to examine the experiences of first-generation individuals, from the applicant stage to the early-career stage. We conducted semi-structured interviews with 41 participants and analyzed responses in accordance with a networked ecological systems theory.

**Results:** The interviews revealed that first-generation individuals put together a patchwork of support. Whereas many MD-PhD trainees struggle at some point in their training, first-generation individuals tend to lack a support system that may provide proactive advice and prepare them for milestones. Interviews shared a common sentiment of isolation due to both a lack of social capital within medicine and academia, as well as a growing disconnect from their families and community.

**Discussion:** Key interventions that would support first-generation students include greater access to high-quality information about the pathway, tailored mentorship throughout training, and more financial resources at the application stage. Trainees and early career physician-scientists seek more flexibility, opportunities for finding community, financial guidance and options, and mentorship around building their careers.

## Introduction

It is widely accepted that a diverse biomedical workforce improves patient care outcomes and increases the impact of scientific research.^1,2^ The importance of diversity pertains not only to biomedical research; studies on corporate performance show a positive correlation between racial and ethnic diversity and a company’s financial returns.^3^ Indeed, the National Institutes of Health has identified diversifying the biomedical workforce as a priority; this includes increasing the representation and support for individuals from disadvantaged backgrounds, including those who “are underrepresented in scientific careers, and have limited access to necessary science and math prerequisites at every academic level”.^4^

Individuals who are the first in their family to pursue higher education, often referred to as first-generation students, are underrepresented in medical school, constituting approximately 11% of matriculants from 2020-2024 despite over two-thirds of the US population not holding Bachelor’s degree.^5^ This is true for dual degree programs as well: 11% of applicants and 8% of matriculants to MD-PhD programs 2021 were first-generation.^6,7^ Aside from underrepresentation, this is an issue because the number of applicants to MD-PhD programs has varied little, despite an almost 20% increase/year in the number of applicants to medical schools from 2012 to 2020. Given that the traditional applicant pool seems to have maxed out, any expansion of the MD-PhD applicant pool will depend on attracting individuals who have been historically under-represented in biomedical research.

Whereas studies have focused on the experiences of first-generation individuals who pursue medical school, none have focused on the challenges that first-generation individuals face in pursuing the physician-scientist pathway.^8–11^ This information is required in order to provide transparent resources and targeted support to aspiring first-generation physician-scientists. One of the challenges in understanding the experiences of first-generation individuals is the intersectionality of their first-generation identity with other factors (e.g., race/ethnicity, gender, sexual orientation, socioeconomic status). Thus, in order to better understand the obstacles first-generation students face, it is important to hear from first-generation individuals themselves.

To fill this knowledge gap, we undertook a phenomenological approach by conducting focus groups with 41 first-generation individuals at different stages of the physician-scientist pathway, including potential applicants, trainees at various stages (students, residents, fellows), and early career physician-scientists. In this study, we compared the experiences of those who were first in their family to pursue a four-year college degree (first-generation) to those who were the first in their family to pursue a graduate degree (continuing-generation). Using this approach, we aimed to identify experiences unique to being first-generation compared to individuals whose parents had access to undergraduate education but did not attend medical or graduate school.

The participants’ described experiences enabled us to identify numerous barriers for first-generation individuals who are pursuing the physician-scientist pathway. Most importantly, participants reported feeling isolated and without sufficient resources at each stage of their training—problems that arose largely due to deficits in access to information and a patchwork of support. Our findings identify interventions to create and bolster access in order to effectively support first-generation physician-scientists before, during, and after training.

## Methods

### Recruitment strategy

This study took a qualitative description approach to examine how first-generation individuals describe their experiences.^12^ First-generation was defined as individuals who were first in their family to complete studies at a four-year undergraduate institution. This group was compared to participants who had at least one parent who had completed a Bachelor’s degree but none who pursued graduate education (defined as continuing-generation). This study was approved by the Weill Cornell Medicine Institutional Review Board.

Participation was open to individuals in the United States who were undergraduate students interested in or currently applying to an MD-PhD or DO-PhD program, students attending an MD-PhD or DO-PhD program, or individuals practicing as physician-scientists who had completed an MD-PhD or DO-PhD program (i.e., resident, fellow, postdoctoral associate, or early career). Information about the study and how to participate was shared via email; information was provided to the American Physician Scientists Association, 115 MD-PhD programs, and 204 physician-scientist or research-oriented residency training programs with a request to share with their members and alumni.

### Data collection

Nine focus groups with a total of 41 participants were conducted by authors BC and BM. The focus groups were organized by identity (first-or continuing-generation and training stage: two for applicants; five for MD-PhD or MD-DO program trainees; one for residents and fellows; and one for early-career physician-scientists. **Table 1** lists sociodemographic information about the participants.

**Table 1.** Participant demographics.

|  | n | % |
| --- | --- | --- |
| Total participants | 41 | 100 |
| Training Stage |  |  |
| Prospective applicants and applicants | 7 | 17 |
| Current trainees | 29 | 71 |
| Preclinical (Pre-PhD) | 8 | 28 |
| Graduate (PhD) | 18 | 62 |
| Clinical (Post-PhD) | 3 | 10 |
| Residents or Fellows | 2 | 5 |
| Early-career | 4 | 10 |
| Race and ethnicity* |  |  |
| *\* Does not add up to 100% because more than one category could be selected* |  |  |
| American Indian or Native American | 1 | 2 |
| Asian | 9 | 22 |
| Black or African American | 5 | 12 |
| Hispanic, Latinx/a/o/e | 7 | 17 |
| Middle Eastern or North African | 1 | 2 |
| White | 25 | 61 |
| Gender |  |  |
| Man | 17 | 41 |
| Woman | 24 | 59 |
| Average yearly childhood household income |  |  |
| < \$25,000 | 9 | 22 |
| \$25,000 - 49,999 | 13 | 32 |
| \$50,000 - 74,999 | 9 | 22 |
| \$75,000 - 99,999 | 7 | 17 |
| > \$100,000 | 3 | 7 |
| First-generation | 31 | 76 |
| Continuing-generation | 10 | 24 |

Focus groups were conducted via Zoom video communications for 1.5 hours using a standardized interview guide developed by BC and BM Focus groups were initially transcribed using Otter.ai and edited for accuracy by authors BC and BM. Themes were identified and agreed on by BC and BM using three randomly chosen transcripts; these included finances, community and connections, work-life balance, access to information, and perceptions and expectations of the physician-scientist career path. Coding for themes was done by authors BC, BM, JW, MCG, RK, and CS with each transcript being reviewed by two authors until concordance was reached.

### Data analysis

A networked version of Bronfenbrenner’s ecological theory of human development was used to analyze the data.^13–16^ Bronfenbrenner’s theory posits that development of an individual is supported by an array of support systems (known as ecological systems) that are nested. The microsystem includes those spaces with which an individual is directly interacting and a part of. Mesosystems encompass the interaction between two of the microsystems. Finally, the exosystem exerts influence on the micro– and mesosystems that an individual belongs to even though the individual is not directly a part of it. A networked version of this theory has been proposed, where the systems interact with each other in a non-hierarchical manner.^13^ We adapted this conceptualization using the themes identified by the study team from the interview transcripts (Figure 1). The first-generation participants were considered to be the central individuals, who interacted directly with individuals within three microsystems: personal support (i.e. friends and family), undergraduate/post-baccalaureate institution (i.e. mentors, faculty, and advisors), and MD-PhD institutions (i.e. advisors, faculty, and training program). Beliefs about the application process were a mesosystem (societal interaction) connecting pre-medical advisors and training programs. The exosystem (setting without the participant) was defined as the culture maintained by MD-PhD training programs, faculty, and practicing physician-scientists.

**Figure 1.**
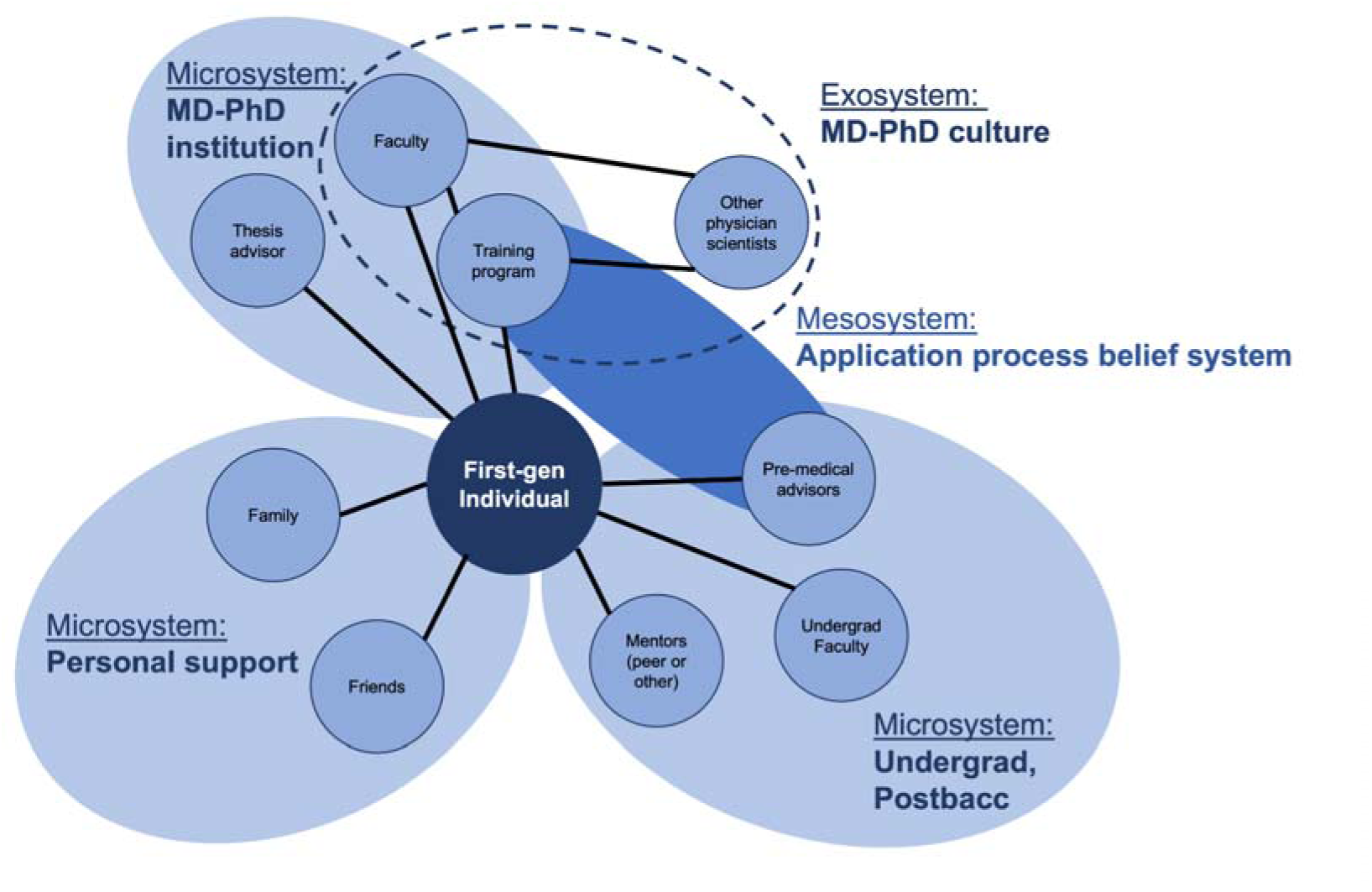
Networked ecological systems theory. Figure illustrates the modified networked EST model used for this study.

**Figure 2.**
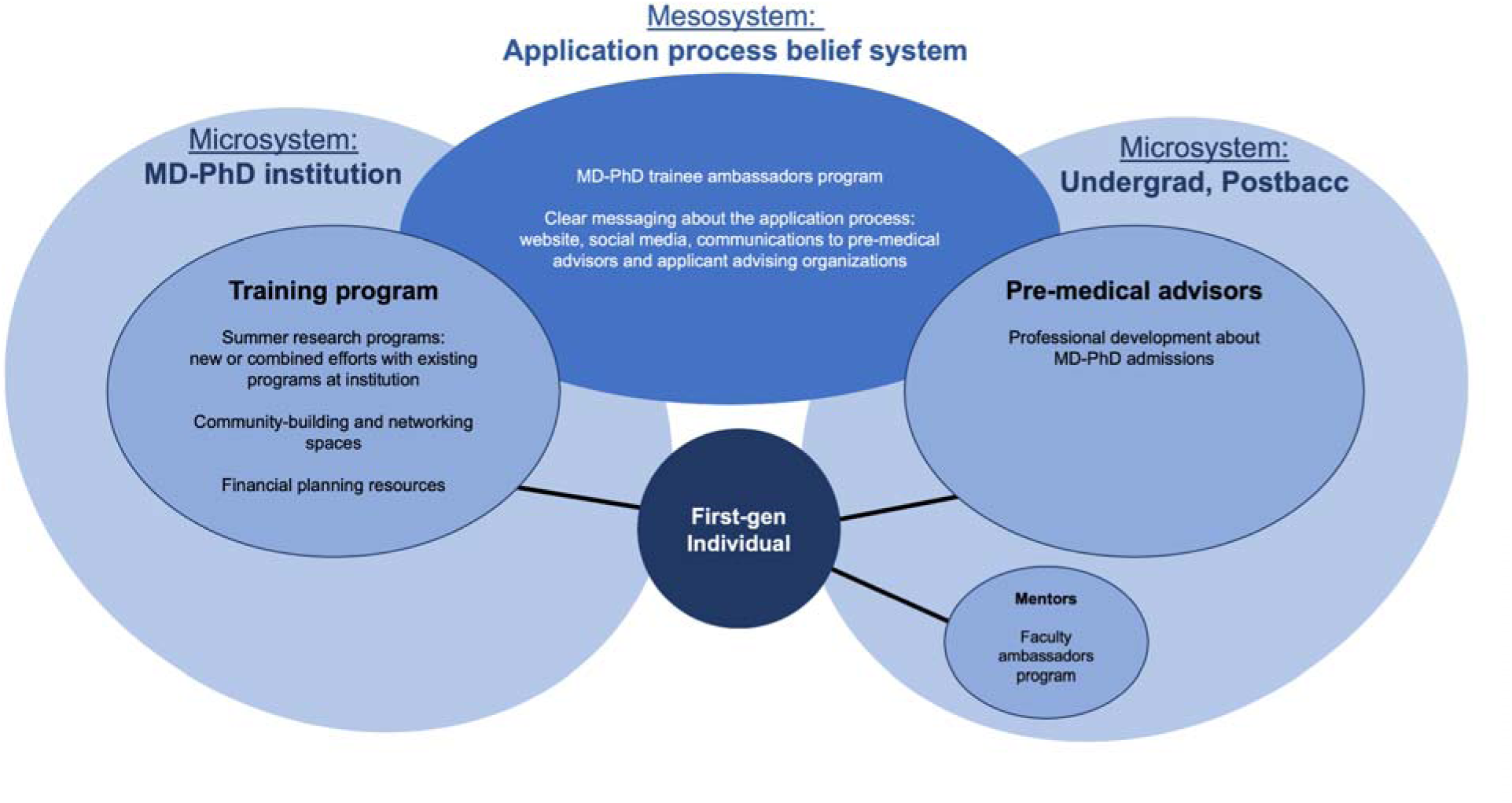
Strategies to support first-generation individuals applying to MD-PhD programs. Figure illustrates how two microsystems (MD-PhD institution and Undergrad/Postbaccalaureate institution) can influence the application process belief system mesosystem to better support first-generation applicants.

## Results

A total of 41 participants were interviewed across ten focus groups: 7 (17%) individuals considering or currently applying to MD-PhD programs, 29 (71%) current MD-PhD trainees, 2 (5%) in residency or fellowship training, and 4 (10%) early career physician-scientists.

Of those interviewed, 31 (76%) were first-generation and 10 (24%) were continuing-generation. In listening to the experiences of both first-generation and continuing-generation interviewees, a common sentiment among all was isolation while pursuing the physician-scientist pathway. These feelings were exacerbated for first-generation individuals, who felt that they lacked access to a solid support network.

### Cultural Barriers

A recurring and predominant theme for first-generation individuals at all stages of training was a clash between the culture they grew up in and expectations of their family with the individuals and environments they encountered in pursuing physician-scientist training. This created a dynamic where first-generation individuals felt out of place both at home and in their current environment, resulting in feeling “split” between the cultures and expectations of both groups. This split identity and lack of central home base enhanced feelings of loneliness and isolation. These feelings were much less likely to arise for continuing-generation interviewees because their family was more likely to understand and even embrace the culture of higher education.

> “I’ve kind of moved into this ivory tower away from my family who is mostly working class … it just feels like I’m kind of the black sheep now to pursue higher education … [but also] it’s hard to fit in with a lot of my medical school peers, who … come from very wealthy backgrounds. So, it’s … this purgatory where I’ve left the community I came from, but I haven’t quite assimilated to this one that I’m joining.”
>
> (Current trainee)

> “It’s really hard, I think, to explain to people back in my community where education is much more about dollars and cents, and about making a living for yourself and for your family. And so, I think this path … has been kind of isolating in that way, in that I not only have to do something that is incomprehensible, but also for reasons that are incomprehensible.”
>
> (Current trainee)

Interviews suggested that isolation manifests in different ways depending on training stage. Prospective and current applicants articulated that sources of isolation included a lack of information and guidance about the pathway, the financial pressures of applying, and an insufficient understanding of the physician-scientist pathway amongst their family members. Participants who were in training or early in their careers described isolation as they navigated the pressures of their financial future and fitting into academia.

### Accessing information about the pathway

Participants noted that they felt that entry into the physician-scientist pathway was especially challenging and often opaque for first-generation individuals, which can discourage many from pursuing this pathway. The lack of readily accessible resources and tailored mentorship, combined with the financial burdens of applying, were noted sources of frustration for many first-generation interviewees. Continuing-generation interviewees experienced overall better support, most likely owing to their families’ personal experiences with higher education (even if not graduate school), as well as the social networks that collective familial education can provide.

At the application stage, first-generation participants reported having to figure out much of the process on their own. This was both because they were less aware of available resources and were not sure how to access ones that were available. Continuing-generation students seemed more likely to be aware of requirements (e.g., significant research experience), resources, and mentorship. Some first-generation individuals participated in summer research programs or other pathway programs that addressed these issues, which highlights a critical need for such programs. Participation in pathway programs gave first-generation individuals more insight into the process and career path; however, many still found it challenging to stay the course when ultimately applying (i.e., “leaky pipeline”).

> “It shouldn’t be this hard [to apply to MD-PhD programs], especially if we want to attract people who are first-generation, you bring in a new set of ideas and perspectives to medicine. We want people like us to be here, and so we shouldn’t make it this hard.”
>
> (Current trainee)

Many participants reported preparing to enter the physician-scientist pathway relatively late compared to their peers. While their continuing-generation peers may have known the steps they needed to take to effectively prepare when they first started college, first-generation individuals depended on mentors or other contacts to inform them about the pathway, which may have occurred much later in their education. As a result, some learned of the physician-scientist pathway during their senior year of college or even during gap years after college, and they had to take more time off between college and the beginning of an MD-PhD program.

> “I kind of came around to research a little bit late … it was something that I never considered because … growing up, I didn’t know any physicians, I didn’t know any scientists.”
>
> (Current trainee)

Most first-generation individuals learned about dual-degree programs through mentors: often professors, coworkers, or lab mates. These mentors were critical to providing information about career options to first-generation students, who may not have personal mentors through their social network. Several interviewees mentioned being involved in high school programs aimed at first-generation individuals that introduced them to research early on through shadowing and hands-on experience. Again, this highlights the importance of pathway programs, especially those that start *before* college.

The earlier that students find out about the breadth of career options available to them, the sooner that they can begin pursuing a particular path. With the research experience required to matriculate into dual-degree programs, it is essential that applicants are made aware of these expectations early on so that they can begin to meet them. Mentorship programs and research opportunities meant to address this need should be openly accessible to all and targeted towards a diverse demographic.

> “My first exposure to research happened in my senior year of high school. I was in a program geared towards first-generation people from low socioeconomic backgrounds… One of the tutors in the program was an undergraduate and was in a lab. I got to shadow him and learn what basic research was all about. And then I ended up joining that lab as an undergraduate again, through that scholarship program that I was in. And so through that scholarship program, I was able to stay in that lab all four years …”
>
> (Current resident or fellow)

Once first-generation individuals were made aware of the dual-degree pathway, many participants experienced challenges in figuring out how to prepare a strong application. First-generation participants often found information online, much of it developed by students who had faced some of the same obstacles. In contrast, continuing-generation interviewees more often cited their mentors or social networks as providing information, although several stated they did also have to search for information online.

A major theme was that many first-generation participants reported feeling discouraged from applying to MD-PhD programs by pre-medical school advisors, who often provided very little information about the dual-degree pathway and in some cases even actively discouraged their advisees to apply. This issue was expressed broadly by all interviewees; however, for first-generation individuals, these advisors were often the only source of information beyond what they found online. Therefore, they tended to be much more reliant on the advice and resources of their pre-medical advisors, as compared to students who had parents who have attended college and have experience with the process of applying to higher education (albeit at the Bachelor’s level) or larger social networks of individuals who had pursued graduate degrees.

One reported reason for this may be that some schools do not have many students applying to dual-degree programs, so their pre-medical school advisors may not have experience preparing students exploring this pathway. In addition to online resources, some participants sought out mentorship from outside organizations, current MD-PhD trainees, or older undergraduate students. This highlights the importance of outreach and education through programs like these.

> “I find it more helpful to look for an outside organization that offers premed mentoring and also advising because that actually provides a wider aspect that’s not centered only around my university. But also it is more centralized around my specific needs and concerns when it comes to the premedical application.”
>
> (Current applicant)

### Affording the application process

Financial pressures made applying to dual-degree programs challenging for many first-generation participants. The majority (65%, n = 20) grew up in families with an annual household income of less than $50,000. As a result, many had to work multiple jobs during college, which meant less time spent studying and participating in extracurricular activities or research—all of which could impact the strength and quality of their application. In addition to raising money for medical school applications, they also faced pressures to finance their education and to support their family. Managing jobs, school, and familial commitments became a huge burden on many first-generation students, especially because many research positions were unpaid.

> “I went to a large R1 research institution, so if you were interested in the sciences…you were told to find labs early…I actually found that experience of trying to find a lab really kind of disheartening and that [principal investigators] wanted you to commit 15 to 20 plus hours a week [yet] weren’t planning on paying you, which was a lot for me…”
>
> (Current trainee)

Not surprisingly, some participants identified work study or other funded research opportunities as making it possible to do research because it alleviated some of the financial pressure, and many reported that they had to apply to a smaller number of programs due to financial constraints. Some took time off after college to not only gain more research experience but also to make money to cover the costs of the application process. Interviewees reported that any financial support that dual-degree programs offered during the application process made a significant difference in their ability to apply.

> “Funding everything independently, myself, I remember [the cost of applying] being the most prohibitive part of applying for MD-PhD programs.”
>
> (Resident/fellow)

> “I ended up having to use all the vacation days I had accrued for about two years to go to interviews…I was limited financially by how many places I could actually afford to travel to pay for all the travel expenses. Fortunately, a lot of that was offset by some of the programs.”
>
> (Current trainee)

Whereas the time commitment involved in interviewing is a problem for all applicants, the financial burden becomes a major issue for first-generation students, who tend to have less resources.

### Isolation during training

#### Justifying the pathway

Dual-degree training is long and, paradoxically, occurs during years when most individuals are experiencing many life transitions that are common during early adulthood (e.g., moving to a new city, supporting oneself, meeting a partner, having children). Participants brought up many challenges related to building a life during and beyond physician-scientist training.

Conversations about finances highlighted a salient paradox: pursuing a physician-scientist career is a financial sacrifice in the long-term, but choosing to attend a (usually funded) dual-degree program made pursuing medicine and research a possibility for those who could not otherwise afford the cost of medical school attendance. First-generation interviewees underscored that the financial support outweighed the risk of taking additional time to prepare to pursue a research career. Becoming a physician-scientist with an MD-only degree was considered out of reach for many participants due to the cost of medical education. There was a large concern amongst participants about taking out loans for their education, especially because many participants had financed their undergraduate degrees through loans that they were already concerned about repaying. Some interviewees mentioned that the financial support for their education meant they did not place an additional burden on their family, even though it also meant that they were not able to contribute financially to their family’s needs while in training.

> “Another major advantage was, I was able to be financially kind of independent from my family, because the program was funded, and I wouldn’t be incurring debt.”
>
> (Current trainee)

However, there was a tension between the perceived financial security of the funded dual-degree training and concerns about earning potential in the future. These concerns were further amplified by family perceptions regarding present and future earnings. The majority of participants felt that their families did not understand what they were training for or assumed that they were already practicing physicians after so much time in school. Whereas continuing-generation peers indicated that their families understood the value of education, first-generation individuals were concerned about their families not understanding why so much further education and training was necessary. First-generation participants felt that their family expected them to provide immediate financial support given the fact that they would be physicians, which is perceived to be a financially-lucrative career path.

Current practicing physician-scientists reflected on lost years of practicing medicine due to the prolonged time-to-degree, resulting in the loss of several years of salary and savings. First-generation interviewees shared that they were unfamiliar with the details of financial planning because this was not information they had received guidance about prior to or during their training years. This “financial penalty” may be less of a concern for more middle– or upper-class families. As a result of the financial pressure accumulated by many years of training, some first-generation individuals put their research pursuits on pause and practiced clinical medicine during the early career phase in efforts to make more money to support their family, which is not a desirable outcome for a dual-degree program.

> “I deviated into medicine solely and have come back to research a little bit later than others because I was able to get that financial stability under my belt from working clinically exclusively for about four years.”
>
> (Early career physician-scientist)

Many interviewees expressed difficulty in communicating with their family about the nature, benefits, and challenges of the physician-scientist pathway. This increased feelings of isolation as trainees could not fall back on their families for support and understanding when they experienced roadblocks and hardships in their programs. Both first-generation and continuing-generation interviewees shared these feelings; however, first-generation individuals experienced having to bridge a larger gap of understanding with family members who had never experienced higher education at any level.

> “Trying to explain what we do and what we’re doing [to] family members … [but] their understanding just stops, like, no matter how hard I try … it’s hard to commiserate when the program is so difficult, especially the PhD portion … [which is] a lot more difficult to portray that … my productivity is directed at these abstract things called papers and presentations and things that don’t have actual meaning or value to people that don’t understand it.”
>
> (Current trainee)

#### Fitting into academia

There are many transitions during physician-scientist training, and academia (especially medicine) is still a very hierarchical environment. Once participants entered dual-degree programs, many struggled with meeting the expectations or milestones of their programs as they navigated making progress during medical and graduate school. First-generation individuals expressed feeling “behind” or “out of place” in academia and struggled to prepare for medical school coursework or navigate finding a supportive lab environment while completing their PhD. Interviewees mentioned how the sense of isolation heightened feelings of imposter syndrome, particularly during the graduate school stage of training. Many did not know where to go for mentorship or guidance because they often could not relate to their PhD thesis advisor.

> “I’ve found a lot of inspiration just from hearing from older students that are further along in the process than me that are so willing to share kind of like what they’re going through right now and what their plans are for the future. I’ve found that there are so many possibilities that I had no idea about, also, not having anyone in the family that’s ever done anything like this.”
>
> (Current trainee)

Prior research experience, especially during gap years, helped to provide a framework for what it meant to earn a PhD. They also learned about what would happen at later stages from their peers.

Many first-generation participants felt disconnected from their peers, who they felt had come from backgrounds very different than their own. First-generation interviewees felt that they did not have community at their institution because they could not relate to the experiences of other trainees, especially regarding wealth or lived experiences. Many described conversations where they realized that being first-generation is not common in medical school and in dual-degree programs.

> “I don’t necessarily share with a lot of classmates just because a lot of them come from very affluent backgrounds, and it gives me kind of the feeling that I’m a little isolated from a lot of them … when they talk about their childhoods, or their families, it just feels so different from what I grew up with.”
>
> (Current trainee)

> “I recall one time … when people were comparing … how many generations [of physicians were in their families]. Some of my classmates … were in their third or fourth generation … I would feel a little bit alienated from my classmates or feeling like I was definitely kind of the odd one out.”
>
> (Resident/fellow)

Interviewees mentioned how the sense of isolation heightened feelings of imposter syndrome, particularly during the graduate school stage of training. Many did not know where to go for mentorship or guidance because they often could not relate to their PhD thesis advisor. Prior research experience, especially during gap years, helped to provide a framework for what it meant to earn a PhD. They also learned about what would happen at later stages from their peers.

> “I’ve found a lot of inspiration just from hearing from older students that are further along in the process than me that are so willing to share kind of like what they’re going through right now and what their plans are for the future. I’ve found that there are so many possibilities that I had no idea about, also, not having anyone in the family that’s ever done anything like this.”
>
> (Current trainee)

Participants noted that it was difficult to find fellow first-generation colleagues at their institution, both because there was a paucity of such students, but also because first-generation status is rarely discussed.

> “I feel like there are certain parts of identity that you can see or you can hear … however, being first-gen is something that doesn’t appear as easily or it’s not something that’s obvious to people.”
>
> (Current trainee)

It was in this context encouraging that supporting first-generation students can act as a positive, reinforcing cycle. Those who have been granted opportunities and mentorship want to pay that forward to the next generation, leading to exponential growth in access.

> “I think a really important part of my career is going to be diversity and inclusion in academic medicine … I hope to run some sort of program, like many of us have participated in, or be a mentor or do something to help people in future generations.”
>
> (Current trainee)

> “The opportunity to work with students and try and help them have a positive experience and pursue their goals and change the world.”
>
> (Current trainee)

## Discussion

Our study highlights that the lack of accessible information about the dual-degree pathway, patchwork of support systems, financial pressures, and familial/external expectations are prevalent factors affecting access to, attrition from, and career satisfaction in the physician-scientist pathway. Many of our study participants shared these stresses as dual-degree trainees in general; however, these stressors were compounded for first-generation individuals.

First-generation participants experience unique challenges that exacerbate common issues for all dual-degree applicants and trainees. Prior to applying, participants noted that they lacked information about what the physician-scientist pathway looked like and how they could best prepare for both the application process and the training itself. They also lacked a support system that could guide them in accessing resources. During their training years, participants reported feeling increasingly isolated from their peers due to their unique backgrounds.

Furthermore, the structure of academia itself felt especially challenging due to their nontraditional backgrounds, including financial struggles and familial commitments. Exacerbating all of this, first-generation students found it not only challenging to fit into academia, but they also felt more disconnected from their communities back at home because their pathways diverged from those of their families and close friends.

In summary, the physician-scientist pathway for first-generation applicants and trainees is fraught with isolation and pressures external to the existing pressures of pursuing a physician-scientist career. In this context, it is essential to understand the first-generation experience and create environments where all students can thrive and reach their full potential. Using the networked ecological systems theory framework, we can identify areas to enhance the network of support for first-generation individuals.

### Prospective and current applicants

There are numerous initiatives that can target the social capital–that is access to social networks–that first-generation individuals may lack.^11,17^ Further support for those applying to MD-PhD programs could come from the baccalaureate microsystem and the MD-PhD program microsystem and, in turn, the application process belief mesosystem perpetuated by the two. Many participants noted that they learned about the MD-PhD pathway from professors and current MD-PhD trainees whom they met during undergraduate or post-baccaulaureate education. Professors and trainees could be recruited as ambassadors by premedical advisors and MD-PhD programs to perform outreach and deliver information to potential applicants; this would require making sure that these ambassadors had up to date information about MD-PhD training in order to make this form of outreach productive. Mentors and information from outside organizations can also bolster the support network for first-generation individuals and provide advice about applying. For example, one participant indicated that a visit by an MD-PhD program director to their college influenced their decision to pursue a physician-scientist career. Some additional examples of organizations that may play a role include the American Physician Scientists Association, the National First-Generation in Medicine Association, MiMentor, Chasing Medicine, MD Collective, and the Program for Underrepresented Medical Applicants Initiative.^18–23^

Additionally, a number of interviewees noted that their first research experiences were summer research programs. Increasing access to research experiences for first-generation individuals will be important since first-generation individuals have significantly fewer research experiences and publications at the time of applying to MD-PhD programs compared to continuing-generation peers.^24^ Prior research at the high school level has shown that afterschool programs can be key in helping students proactively learn about requirements for pursuing a college education; a similar approach could be taken to introduce more resources and support for first-generation participants.^25^ More MD-PhD programs could sponsor or partner with existing undergraduate or post-baccalaureate research programs at their institutions, similar to the model of the Gateways to the Laboratory Program, to tailor programs to first-generation students.^26^ As part of the Gateways program, undergraduates work in a research lab while also receiving targeted mentorship and professional development from MD-PhD students and practicing physician-scientists. It is important that these programs be appropriately funded to ensure that students do not have to choose between research opportunities and earning money during the summer. Virtual options could also be explored, such as the Virtual Summer Research Program established by the American Physician Scientists Association.^27^

A common theme in our study was a lack of encouragement to pursue the MD-PhD pathway, especially from pre-medical advisors. Clearer messaging to interested students and pre-medical advisors from programs regarding their requirements and goals could be helpful in addressing the belief that first-generation students may not be competitive applicants.^8^ The interviewees in our study identified websites and online resources as critical to their approach in applying to programs. A previous study of medical students from underrepresented minority backgrounds further emphasizes this by revealing that they did not pursue the MD-PhD pathway because there was a dearth of information about what programs are looking for.^28^ Many MD-PhD programs do not include even basic information about statistics like GPA and MCAT scores of applicants and matriculants to their program, which is an opportunity for transparency that would benefit first-generation applicants.^29^

Increasing knowledge and availability of financial resources during the application process was also a stated need by those interviewed in this study. The creation and accessibility of fee waivers and options for interview cost coverages are options to address these needs. Educating families of potential first-generation applicants about the pathway would also provide necessary context for the family to support the individual. For example, in the Gateways to the Laboratory Program, families are invited to the final research symposium (and travel/lodging costs are covered) so that they can not only see the student share their capstone research presentation but also learn about what it means to be a physician-scientist.

### During training

First-generation medical students report higher rates of stress and fatigue in addition to financial concerns, lower quality of life, and social support.^30^ The length of time to complete the dual degree presents some unique challenges for first-generation trainees. Whereas MD-PhD students in general experience moments of isolation during the training, these experiences are compounded for first-generation individuals.^31^ Participants emphasized needing greater support from their families and MD-PhD programs.

Family can be both a positive and a negative for first-generation individuals. The experience of being first-generation can magnify independence as they must navigate the systems of higher education while potentially feeling guilt for leaving their families.^32^ Additionally, individuals often feel that they must constantly prove that the extended training is worth the investment, particularly given the financial prospect of entering medical practice more directly.^33^ A newsletter sent to families similar to My MD-to-Be that is tailored to the MD-PhD experience would be a valuable tool to provide families with context about the milestones a student is working on at that stage of training.^34,35^ MD-PhD programs could do information sessions for families when they may be visiting such as the white coat ceremony.

Being able to contribute financially to their family and build their own nuclear family was also described as challenging given the expectations that being a physician should bring financial resources. The stipend makes the MD-PhD pathway attractive to first-generation individuals because it helps offset the sacrifice of pursuing a research career; previous studies have found that first-generation medical students are more likely to report wanting to pursue loan forgiveness programs, suggesting that finances are a concern in general.^36^ One potential concern is that some individuals might pursue an MD-PhD program as way to go to medical school at no cost without a strong desire to be researchers. However, the increasing prevalence of tuition– or debt-free medical school options assuages this concern because it provides an alternative path for those who were not as interested in research from the start. Financial advising and support can be provided by MD-PhD programs to benefit all trainees, but in particular first-generation students as they navigate their personal responsibilities with fixed financial resources. Some areas where better support could be built are relocation expenses at the start of the program, childcare options, healthcare for dependents, accommodations for illness/disability, paid parental leave, residency application costs, loan forgiveness, and emergency funds.

Participants underscored feelings of isolation within the medical school and academia, which affected their ability to build a career. This is a clear opening for MD-PhD programs and institutions to support first-generation individuals as trainees and practicing physicians. The cultural mismatch (exosystem) is in part due to having different experiences, such as tastes and hobbies, compared to peers and faculty.^37^

First-generation individuals may also benefit from guidance on networking when they enter this new space as a way of developing social capital.^11^ Another identified need was the creation of opportunities to connect with other first-generation individuals at various stages of training, which MD-PhD programs could help facilitate. In fact, interviewees at later stages, including those who have completed training, expressed a desire to connect with those at earlier stages. There is an opportunity for programs to encourage first-generation trainees at different stages to come together and share their experiences and advice as a form of peer mentorship.

Mentorship needs to be attuned to the needs of first-generation individuals. In describing professional development, interviewees described uncertainty about building their careers due to a lack of knowledge about next steps. Mentorship could help first-generation physician-scientists navigate academic hierarchy and effectively plan the next stages of career development.^38^ Programs and institutions that employ first-generation physician-scientists can develop connections and community amongst these individuals; this would help ensure the success of first-generation individuals, while also providing resources that would dismantle the hidden curriculum of academic medicine. For example, institutions could incorporate goal setting and transparent conversations about available resources available into annual evaluations and individual development plans. The Association of American Medical Colleges has compiled tools for advisors aimed at supporting first-generation individuals that could be adapted to later training stages as well.^39,40^ Future studies should examine the experiences of first-generation physician-scientists currently in the workforce.

This study has limitations, including selection bias. Our sample was derived from those who indicated interest in participating in the study after having received the solicitation sent out by partners, and thus self-selection bias and bias in outreach must be recognized. Our focus groups did not include individuals whose parents had advanced degrees in medicine and/or science. However, we used individuals who were first in their family to pursue a graduate degree or higher education in the US as a sort of internal control for those who might have more or different information about navigating higher education. Additionally, the small number of physician-scientists at later career stages (resident and beyond) restricts the ability to identify other challenges experienced beyond training. The interviewees therefore may not be fully representative of all first-generation individuals interested in and who trained at US MD-PhD programs.

## Conclusion

This study is the first to examine the experiences of first-generation physician-scientists in training and early career. The common theme of isolation was articulated by many participants, emphasizing the need for greater action to bolster the various support networks that would benefit first-generation individuals. Many interventions would also benefit continuing-generation applicants and trainees since they would provide more transparency and mentorship overall. Including individuals with a diversity of experiences is important for growing a workforce not only representative of the US population but also for incorporating different perspectives to improve medicine and research. Multi-pronged efforts and commitment by collegiate, postbaccalaureate, MD-PhD training and employing institutions are required to address the pressures faced by those who are first-generation. Future work should continue to be driven by the input of first-generation individuals to inform targeted and productive interventions to serve this community and ensure their success as physician-scientists.

## Ethics statement

This study was deemed exempt from review by the Weill Cornell Medicine institutional review board because it entailed interviews and information was kept deidentified.

## Author contributions

BC and BM contributed to conceptualization, methodology, creation of the interview guide and conducted the interviews. BC, BM, JW, MCG, RK, and CS contributed to coding the interviews and identification of themes. All authors contributed to writing, reviewing and editing the manuscript. OSA and CB supervised the project.

## Data Availability

All data produced in the present study are available upon reasonable request to the authors.

## Acknowledgments

We would like to thank Harlan Pietz for help with initial logistics in planning this study.

